# Automated development of clinical prediction models enables real-time risk stratification with exemplar application to hypoxic-ischaemic encephalopathy

**DOI:** 10.1101/2022.04.01.22273313

**Authors:** Matthew S. Lyon, Heather White, Tom R. Gaunt, Deborah Lawlor, David Odd

## Abstract

Real-time updated risk prediction of disease outcomes could lead to improvements in patient care and better resource management. Established monitoring during pregnancy at antenatal and intrapartum periods could be particularly amenable to benefits of this approach. This proof-of-concept study compared automated and manual prediction modelling approaches using data from the Collaborative Perinatal Project with exemplar application to hypoxic-ischaemic encephalopathy (HIE). Using manually selected predictors identified from previously published studies we obtained high HIE discrimination with logistic regression applied to antenatal only (0.71 AUC [95% CI 0.64-0.77]), antenatal and intrapartum (0.70 AUC [95% CI 0.64-0.77]), and antenatal, intrapartum and birthweight (0.73 AUC [95% CI 0.67-0.79]) data. In parallel, we applied a range of automated modelling methods and found penalised logistic regression had best discrimination and was equivalent to the manual approach but required little human input giving 0.75 AUC for antenatal only (95% CI 0.69, 0.81), 0.70 AUC for antenatal and intrapartum (95% CI 0.63, 0.78), and 0.74 AUC using antenatal, intrapartum, and infant birthweight (95% CI 0.65, 0.81). These results demonstrate the feasibility of developing automated prediction models which could be applied to produce disease risk estimates in real-time. This approach may be especially useful in pregnancy care but could be applied to any disease.

## Introduction

In healthcare, prediction is used to stratify individuals into different groups of risk to optimise prevention and treatment of disease^1^. This approach has been successfully applied for decades to identify individuals who will benefit from statin medication, reducing population levels of cardiovascular disease^2^. Prediction modelling involves combining several variables in a statistical model (for example a multivariable regression model) to estimate the occurrence of a future outcome^1^. Clinical prediction models should be accurate^3^ (i.e., discriminate between those who go on to get the disease and those who remain healthy, and calibrated^3^ such that the proportion predicted to get disease is similar to that subsequently observed), feasible (i.e., using variables that are relatively easy to obtain) and cost-efficient. New prediction models should be compared to those already used in terms of accuracy, feasibility, and cost.

Deciding which variables to include in a prediction model is one of the most important and difficult steps in the model development process^4^. Traditionally, prediction models have been based on established causal risk factors, for example cardiovascular risk prediction tools in common use internationally, such as the pooled cohort equation^5^ and Q-risk^6^, include established cardiovascular risk factors such as cigarette smoking, systolic blood pressure and diabetes. However, identifying risk factors is time consuming and laborious and may be subject to selection biases if these variables were preselected using prior knowledge. This process may not be optimal or practical when applied to large datasets and must be repeated with every new dataset. Furthermore, it is increasingly recognised that whilst some risk factors are good predictors, disease predictors do not need to be (causal) risk factors.

The availability of large multidimensional data such as multi ‘omics and electronic health records (EHRs) has enabled the potential to take a hypothesis-free approach to prediction which may identify more variables that could improve prediction accuracy. Automated feature selection methods are often used in these studies^7–9^ to reduce the potential number of variables that remain in the model. These are needed as the scale of the data (several 100s or 1000s of variables) would be beyond conventional approaches comparing different multivariable regression models with a smaller number of variables selected on the basis of ‘independence’ based on a p-value and/or regression coefficient threshold. Several popular feature selection approaches exist including penalised regression^10,11^, decision tree ensembles^12^ and recursive feature elimination (RFE)^13^, among others.

With the increased use of EHRs and widespread availability of computing resources in primary and secondary care it should be feasible to implement dynamic prediction models^3^. These are tools that enable ‘real-time-updated’ risk stratification for different health outcomes that refresh on each clinic appointment, or as new data are collected so treatment is tailored to that new risk. For example, a recent study demonstrated a pipeline for use with structured EHRs and its application to predicting risk at regular time intervals, such as every 6-hours in patients with acute hospital admissions^14^. In parallel with this, new data from all patients could be used to regularly update the underlying prediction models for different health outcomes. Such ideas would require major changes to the understanding of prediction and risk stratification by the public and healthcare providers, together with major operational changes to healthcare systems and provision. Determining the extent to which automated prediction modelling applied to EHRs could improve prediction of health outcomes over and above established risk factors would be useful for knowing whether the aim of ‘real-time updated prediction’ for ‘personalised’ healthcare is feasible, and if so, for which conditions.

Real-time updated risk stratification could be particularly beneficial in pregnancy care as data are collected at regular intervals during antenatal and intrapartum (i.e., around the time of labour and birth) periods. However, unlike application to acute hospital admissions, pregnancy data are collected by a wide range of healthcare providers leading to rich but highly unstructured data that require robust prediction approaches.

A significant cause of perinatal brain injury is perinatal asphyxia, leading to hypoxic-ischaemic encephalopathy (HIE) affecting 2-6 per 1000 live births^15^. HIE is often devastating, with life-long impacts for the infant^16^ and their family, as well as costing society millions of pounds in medical compensation, lost earningsearnings, fare support^17^. While preventative interventions such as induction of labour or operative delivery can be employed if the risks of continuing the pregnancy are higher than those of early delivery^18^ there is a lack of clear data on when to intervene.

Badawi *et al*^19,20^ identified thirty-five potential (independent) antenatal and intrapartum risk factors for HIE. As these factors were identified individually (using univariate analyses) in a case-control study, the predictive accuracy of models combining these risk factors were not explored in the original studies and to the best of our knowledge predictive accuracy of these factors for HIE has not been explored in publications since then.

The aim of this study is to explore the feasibility of undertaking real-time updated risk stratification in pregnancy using a large linked antenatal and intrapartum dataset applied to prediction of HIE, as an exemplar of the potential of health record clinic data to improve risk prediction. To do this we used data from the large Collaborative Perinatal Project and applied automated feature selection approaches to prepare and evaluate prediction models using antenatal only and antenatal and intrapartum variables. These models were compared with the 35 potential risk factors manually identified by Badawi *et al*.

## Methods

### Participants

The Collaborative Perinatal Project (CPP) is a prospective cohort study of approximately 60,000 pregnancies, and 58,000 live born infants born between 1959 and 1965 throughout the prenatal period, labour, and delivery, postpartum and as the child grew^21^. Participants were removed from analyses if the pregnancy was preterm (< 37 weeks), post-term (> 42 weeks), or of young mothers (< 16 years). CPP data files are freely available for download from the National Archives Catalog (https://www.archives.gov/research/electronic-records/nih.html). Ethical approval was not required for this study as the data were publicly available for unrestricted use as detailed in this note (https://catalog.archives.gov/OpaAPI/media/643929/content/arcmedia/electronic-records/rg-443/CPP/304.1-3ND.pdf).

### Outcome

HIE was defined as having definite seizures, hypertonia, jitteriness, hypotonia, abnormal reflexes, or abnormal cry (provided in the CPP dataset without guidance on clinical criteria used); after having a low 5 minute Apgar score (<7)^22^. Data were collected by independent observers onto dedicated forms independently of the clinical team.

### Defining training and testing datasets

Pregnancies were ordered chronologically and split into two equal subsets for training (infants born 1959-1962) and testing purposes (infants born 1963 to 1965). All variables were categorised as either antenatal (measurable before 37 weeks’ gestation), intrapartum (measures only available at or after 37 weeks, up to the point of delivery), and infant birthweight, and classified as either unordered categorical, ordinal, or numerical. The earlier pregnancies were used for training to emulate the use of existing records in predicting future events.

### Feature preparation

Variables were excluded if they contained >10% of missing data values (n=28/518) in the whole dataset leaving a potential n=490 exposure data fields for the prediction models. Differences in missingness between training and testing datasets were generally low except for private health insurance which was 13.5% missing in the training dataset and 0% in the testing cohort (**Supplementary Figure 1**). Unordered categorical variables were recoded as dummy variables producing n=2126 additional fields (**Figure 2**). Variables with zero variance were removed from analysis (n=189), the remaining features were split into antenatal (n=1638), antenatal and intrapartum (n=2122), and antenatal, intrapartum and birthweight (n=2427). All features were standardised to have zero mean and unit variance.

Broadly, two approaches were used to select features for developing HIE prediction models (**Figure 2**). First, a manually defined set were prepared by taking 20-35 potential risk factors for HIE (**Supplementary Table 1**) identified in Badawi et al^19,20^. Second, automated feature selection approaches applied to training dataset were used to rank features in terms of predictive performance to select the top n=20, n=40 or n=60 predictors for each selection method. The feature selection methods used were logistic regression with RFE, Elastic net and least absolute shrinkage and selection operator (LASSO) regression, random forest classifier, and linear support vector classifier with LASSO regularisation (SVC), as outlined below and in Supplementary Table 2.

#### Reverse feature elimination^13^ with cross-validation

During the first iteration all input variables were included as predictors in a logistic regression model trained using five-fold cross validation. On each subsequent iteration the five weakest predictors (determined by the smallest absolute coefficient) were eliminated, with iterations continuing until only one predictor remained. This approach uses the effect of each predictor on the cross-validation mean AUC to rank features. The presented rankings were inversed so that the metric was positively correlated with feature predictability and to allow between-method comparisons.

#### LASSO^10^ and Elastic net^11^ regression

Logistic regression with L1 (shrinks weak coefficients to zero, LASSO) or L1 & L2 (combined shrinking and removal of weak coefficients, Elastic net) regularisation penalties was trained using five-fold cross-validation to determine the optimal value of alpha. The Elastic net mixing parameter (l1_ratio, representing the ratio of L1 [shrinks weak coefficients to zero] to L2 [removes weak coefficients] penalties) was set to 0.5. The penalty term shrinks weak predictors to zero which were subsequently eliminated from downstream analyses. The remaining features were ranked by their absolute regression coefficient (smallest is least predictive).

#### Random forest classifier^12^

The random forest classifier was trained using five-fold cross-validation and default parameters to estimate the feature importance metric which was used to rank features (smallest is least predictive).

#### Penalised linear support vector classifier^23^

The linear SVC was trained with five-fold cross-validation using the AUC metric. The default L1 penalty term (C=1.0) was applied to shrink weak coefficients to zero. The coefficients were taken as a measure of feature importance (absolute value; smallest is least predictive).

### Binary classification

The primary classification analysis was performed using logistic regression models trained with training data and applied to estimate HIE probability using the testing data. Follow on analyses compared logistic regression with a range of other classifiers using default hyperparameters: random forest, naïve Bayes, and support vector machine (radial basis function kernel) and neural network (one hidden layer with number of nodes equal to number of predictors using the rectified linear activation function and Adam optimiser^24^). Model discrimination was compared using area under the receiver-operator curves (ROC) with 95% confidence interval.

### Software

Data were pre-processed using Stata (v13) and Python (v3.8) with pandas (https://pandas.pydata.org; v1.3.5) and numpy^25^ (v1.21.5) packages. Prediction models were developed using Stata, scikit-learn^26^ (v1.0.2) and Tensorflow (v1.15.0; www.tensorflow.org). R (v4.0.2) was used to estimate AUROC and 95% confidence interval with pROC package^27^ (v1.16.2) and prepare plots with ggplot2^28^ (3.3.3) and GGally (https://github.com/ggobi/ggally; v2.0.0) packages.

## Results

### Participants

The dataset was based on the full CPP variable file dataset, containing data on 58,760 infants (**Figure 1**). A total of 12,005 infants were born preterm (<37 weeks of completed gestation), 5476 were born after 42 weeks, and 964 were born to a mother of younger than 16 years age; leaving a total of 40,315 for the analyses. 19,487 infants were born between 1959 and 1962 (and were placed in the first cohort), while 20,828 were born between 1963 and 1966 (and were placed in the second).

**Figure 1.**
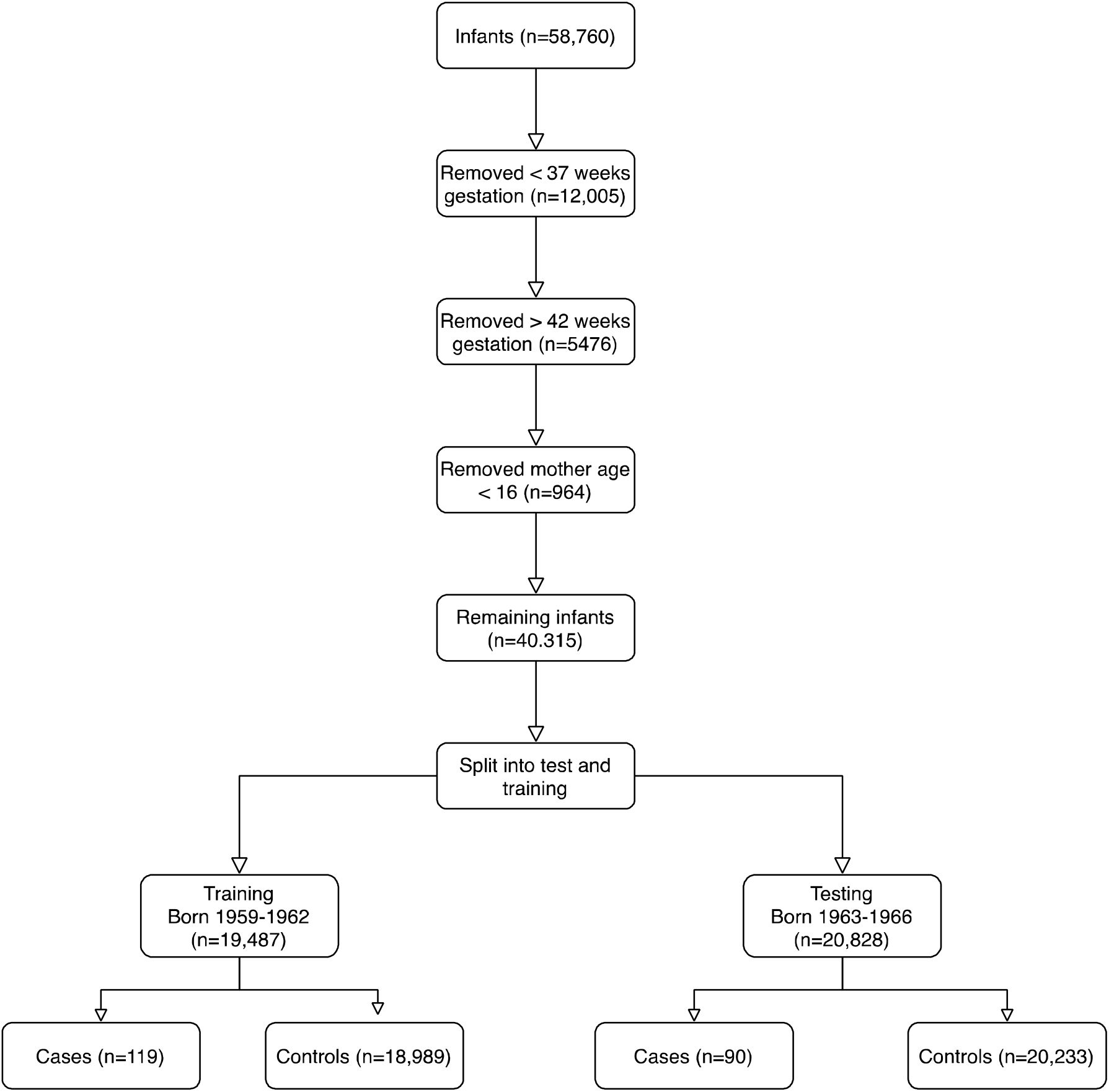
Participant inclusion flowchart.

Table 1 shows characteristics of mothers and infants by HIE status. Overall, 209 (0.5%) had evidence of HIE, 549 (1.4%) died in the perinatal period, 1228 (3.1%) had a low Apgar score at 5 minutes and 2013 (5.1%) required resuscitation after birth. Measures varied by many of the demographic measures recorded, with infants with HIE more likely to have older mothers (e.g., they were less likely to be employed [11.2% vs 15.5%]), have private health insurance (2.7% vs 7.0%) or be of white ethnicity (30.1% vs 49.9%). Mothers were also more likely to have antenatal complications in their pregnancy (e.g., more preeclampsia (9.1% vs 3.3%) and pre-labour bleeding [33.8% vs 28.1%]). Infants with HIE also differed in their birth characteristics (e.g., breech presentation 15.3% vs 2.7%), their sex (62.1% vs 50.6% male) and were smaller.

**Table 1.** Demographics of study population split by HIE.

| Characteristic |  | Non-HIE infants | HIE infants | P |
| --- | --- | --- | --- | --- |
| **Ante-natal Measures** |  |  |  |  |
| Late Booking*** |  | 11,405 (29.1%) | 60 (28.7%) | 0.905 |
| Thyroid Disease |  | 1,028 (2.6%) | 5 (2.4%) | 0.837 |
| Maternal Age | < 20 years | 11,057 (28.2%) | 57 (27.3%) | <0.001 |
|  | 20-24 | 11,690 (29.8%) | 49 (23.4%) |  |
|  | 25-29 | 8809 (22.5%) | 36 (17.2%) |  |
|  | 30-34 | 4,644 (11.8%) | 39 (18.7%) |  |
|  | 35 or more | 3,022 (7.7%) | 28 (13.4%) |  |
| Parity | 0 | 10,434 (26.7%) | 78 (37.3%) | 0.001 |
|  | 1 | 8579 (22.0%) | 29 (13.9%) |  |
|  | 2 or more | 20,049 (51.3%) | 102 (48.8%) |  |
| Employed |  | 5,989 (15.5%) | 23 (11.2%) | 0.084 |
| Private Insurance |  | 2546 (7.0%) | 5 (2.7%) | 0.022 |
| Race | White | 19,560 (49.9%) | 63 (30.1%) | <0.001 |
|  | Black | 16,898 (43.1%) | 123 (58.9%) |  |
|  | Other | 2,764 (7.1%) | 23 (11.0%) |  |
| Family History of Seizures |  | 2560 (6.7%) | 16 (7.9%) | 0.500 |
| Family History of Neurology* |  | 1479 (3.9%) | 12 (5.9%) | 0.133 |
| Fertility Investigations |  | 1008 (2.6%) | 6 (2.9%) | 0.797 |
| Hypertension |  | 167 (0.4%) | 1 (0.5%) | 0.911 |
| Preeclampsia |  | 1284 (3.3%) | 19 (9.1%) | <0.001 |
| Maternal Height | <160cm | 13,221 (36.4%) | 81 (41.3%) | 0.354 |
|  | 160-164cm | 10,961 (30.2%) | 54 (27.6%) |  |
|  | >164cm | 12,172 (33.5%) | 61 (31.1%) |  |
| Pre-labour bleeding |  | 10,792 (28.1%) | 69 (33.8%) | 0.071 |
| Antenatal Viral Illness |  | 2,688 (6.9%) | 15 (7.2%) | 0.846 |
| Alcoholism |  | 44 (0.11%) | 0 (0.0%) | 0.628 |
| Fever |  | 5,068 (13.0%) | 26 (12.4%) | 0.817 |
| Male |  | 19,842 (50.6%) | 134 (62.1%) | <0.001 |
| Placental Previa |  | 160 (0.41%) | 3 (1.5%) | 0.020 |
| Multiple Birth |  | 290 (0.74) | 5 (2.4%) | 0.006 |
| **Intra-partum measures** |  |  |  |  |
| OP presentation |  | 2512 (6.6%) | 34 (16.8%) | <0.001 |
| Breech Presentation |  | 1023 (2.7%) | 31 (15.3%) | <0.001 |
| ROM>12 hours |  | 5706 (16.5%) | 50 (30.5%) | <0.001 |
| Caesarean Section |  | 2076 (5.3%) | 38 (18.2%) | <0.001 |
| MIE** |  | 3000 (7.7%) | 42 (20.1%) | <0.001 |
| Nuchal cord |  | 10,225 (26.3%) | 52 (24.9%) | 0.636 |
| Prolapsed cord |  | 311 (0.8%) | 12 (5.7%) | <0.001 |
| Onset | No Labour | 1,146 (3.0%) | 12 (5.8%) | 0.019 |
|  | Spontaneous | 35,124 (90.3%) | 177 (85.1%) |  |
|  | Induced | 2636 (6.8%) | 19 (9.1%) | 0.019 |
| Shoulder Dystocia |  | 230 (0.6%) | 9 (4.3%) | <0.001 |
| Epidural |  | 617 (1.6%) | 10 (4.9%) | <0.001 |
| **Birthweight** |  |  |  |  |
| Birthweight centile | Less than 3<sup>rd</sup> | 1208 (3.1%) | 29 (14.1%) | <0.001 |
|  | 3<sup>rd</sup> to 10<sup>th</sup> | 2898 (7.4%) | 28 (13.6%) |  |
|  | 10<sup>th</sup> to 90<sup>th</sup> | 31,265 (79.8%) | 125 (60.7%) |  |
|  | Above 90<sup>th</sup> | 3824 (9.8%) | 23 (11.7%) |  |

HIE, hypoxic-ischaemic encephalopathy. FHx, family history. OP, occiput posterior fetal
position. CS, caesarean section. ROM, rupture of membranes.
\* Motor, sensory, or developmental disorder in siblings
\*\* APH, eclampsia, uterine rupture, or ruptured cord
\*\*\* >26 weeks of gestational age

### Automated feature preparation

Of the 518 variables provided in the CPP dataset (**Figure 2**), 28 (5%) were removed due to high missingness (>10%). The variables were split by type into ordinal (n=26), continuous (n=27) and unordered categorical (n=437). Unordered categorical variables were split into n=2563 dummy variables. The combined set were filtered to remove features with zero variance leaving n=2427 features which were split by collection point during pregnancy into antenatal (n=1638), antenatal and intrapartum (n=2122) and antenatal, intrapartum and birthweight (n=2427).

**Figure 2.**
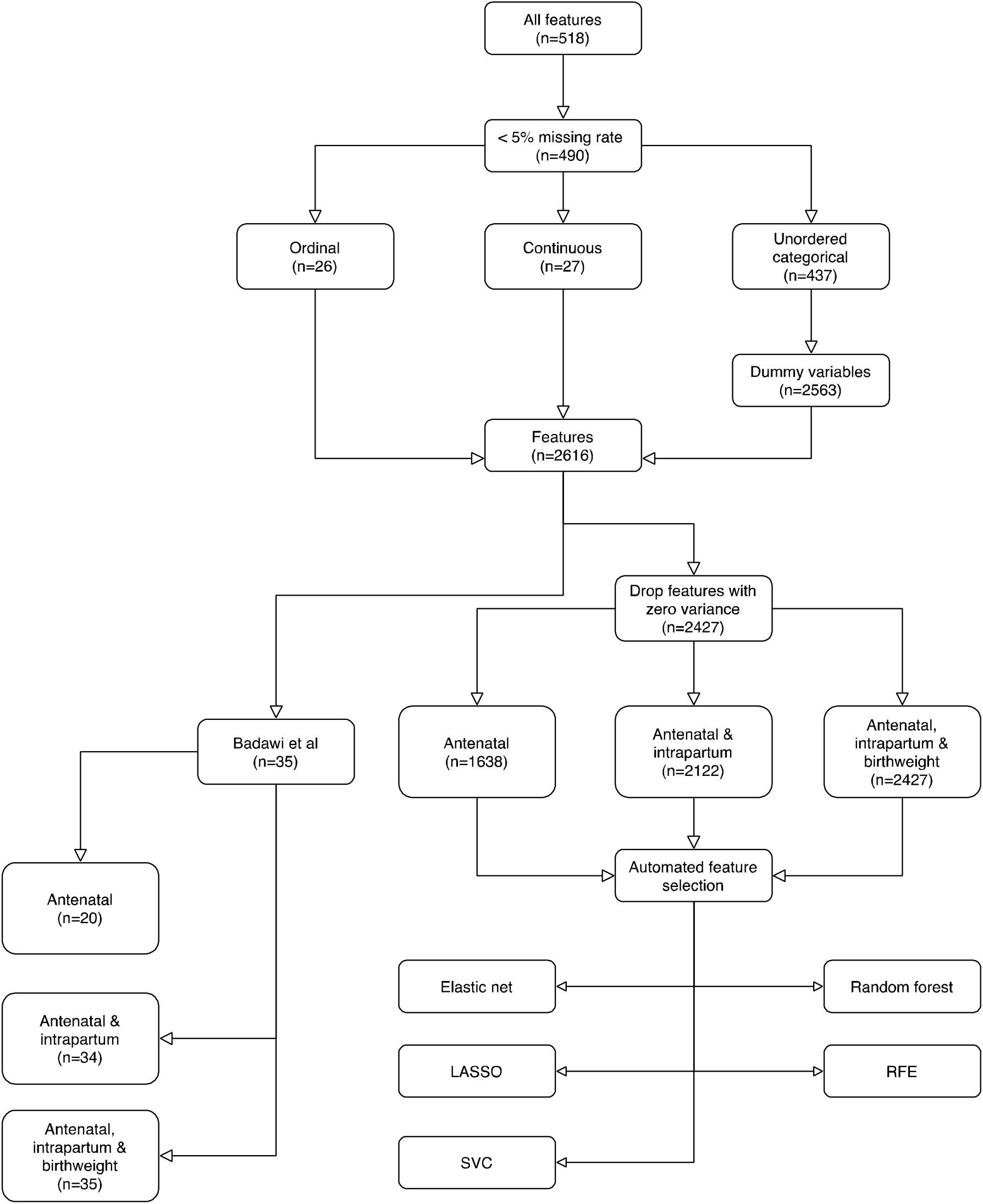
Feature engineering and selection workflow. LASSO, least absolute shrinkage and selection operator. RFE, reverse feature elimination. SVC, penalised linear support vector classification.

Feature rankings for Elastic net, LASSO, penalised linear SVC, and random forest (Spearman’s correlation coefficient 0.23 to 0.71) were most highly correlated, while RFE and LASSO/Elastic net were less well correlated (Spearman’s correlation coefficient 0.17 to 0.34; **Supplementary Figures 2-4**).

### Model performance

The discriminatory ability of each feature set was estimated using the AUC on the second (later) half of pregnancies with logistic regression (**Figure 3**).

**Figure 3.**
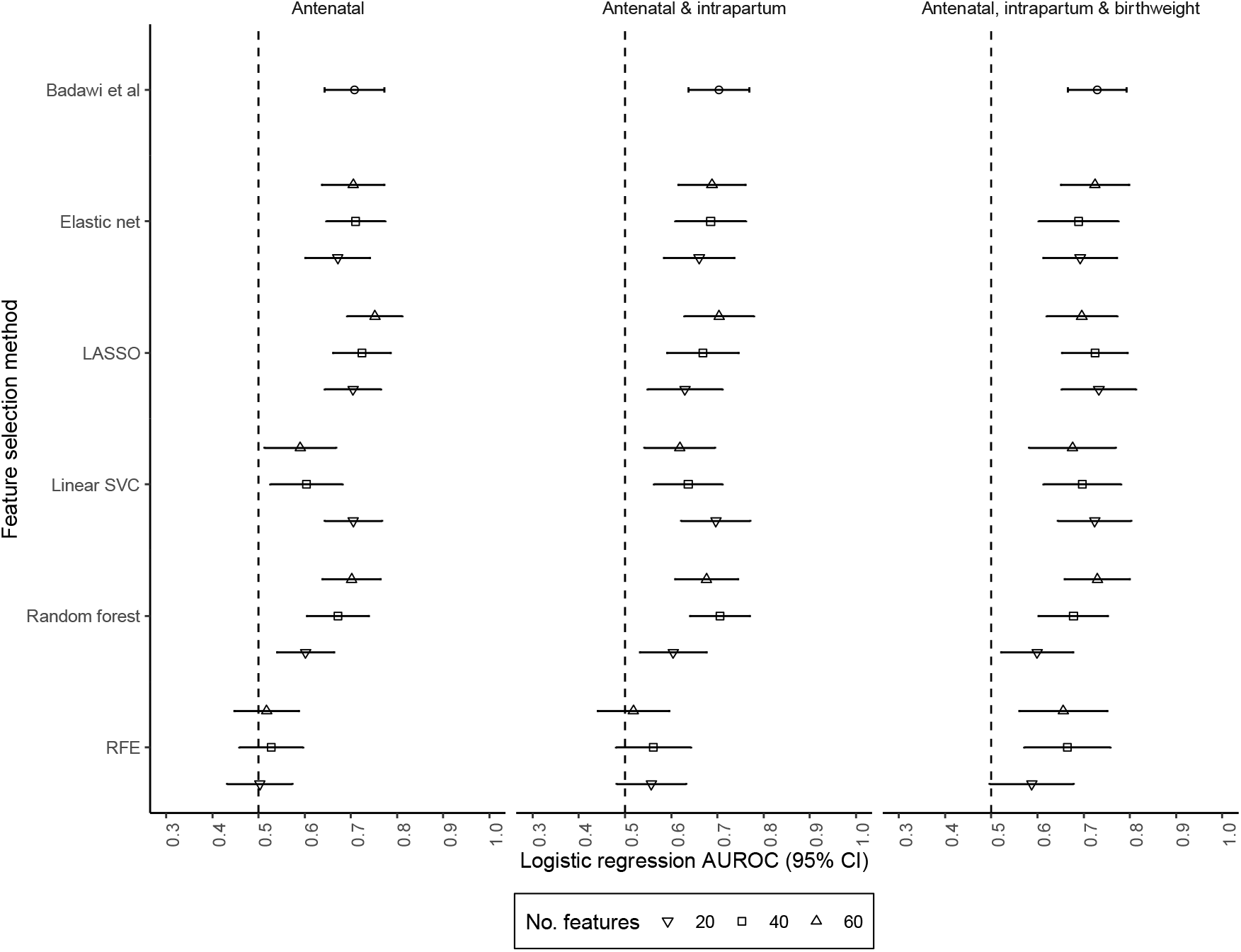
HIE prediction using logistic regression with a range of feature selection approaches. Area under ROC for prediction of hypoxic-ischaemic encephalopathy using logistic regression and a range of feature selection approaches. The model was trained using the first 50% infants (born 1959-1962) and evaluated using the latter 50% of infants (born 1963 to 1965). AUROC, area under the receiver operator curve. CI, confidence interval. LASSO, logistic regression least absolute shrinkage and selection operator. RFE, reverse feature elimination using logistic regression. SVC, penalised linear support vector classification.

Established predictors of HIE gave equally good discrimination using features collected at each of the pregnancy time points. For example, antenatal features gave an AUC of 0.71 (95% CI 0.64-0.77; n=20 predictors), antenatal and intrapartum measures had an AUC 0.70 (95% CI 0.64-0.77; n=34 predictors), and antenatal, intrapartum, and infant birthweight was 0.73 AUC (95% CI 0.67-0.79; n=35 predictors).

The automated feature selection approach was applied to select the top n=20, n=40 or n=60 features using antenatal only, antenatal and intrapartum, and antenatal, intrapartum, and infant birthweight measures and modelled using logistic regression for comparison with established predictors (**Figure 3**). Broadly, there was no strong difference in discrimination between the clinically defined and Elastic net, LASSO, penalised linear SVC, and random forest feature selection approaches. RFE performed worse than the clinically defined features at antenatal and antenatal and intrapartum timepoints. Overall, there was a trend towards better discrimination with a larger number of features included in the model. However, the models derived using features identified by Badawi et al gave better performance than the automated approaches with similar numbers of predictors. Adding intrapartum and birthweight variables had no strong impact on discrimination.

There was no strong improvement in discrimination using a range of classifiers in comparison with logistic regression (**Supplementary Figure 5**).

## Discussion

Here we demonstrate the feasibility of developing automated and low-cost prediction models with minimal human input as a proof of concept for providing real-time updated patient risk stratification which could be applied to any disease outcome. As an exemplar this approach was applied to classify HIE using variables collected at different timepoints throughout pregnancy and benchmarked with manually prepared models.

Automated development of logistic regression models using features ranked by LASSO gave best performance of all automated feature selection methods. These were 0.75 AUC (95% CI 0.69, 0.81) using antenatal only, 0.70 AUC (95% CI 0.63, 0.78) for antenatal and intrapartum and 0.73 AUC (95% CI 0.65, 0.81) for antenatal, intrapartum, and infant birthweight which were not substantially different from logistic regression models developed using features selected manually by Badawi *et al*. Meanwhile, feature ranking with RFE produced models with lower discriminative capacity and this approach may not be suitable for providing real-time updated risk estimation. Our results demonstrate the utility of automated feature selection approaches in developing accurate risk prediction models. This process could be optimised to produce models with little or no human input and updated in real-time as new data are made available. This is in contrast with the manual approach taken by Badawi et al which is time-consuming and laborious, requires expert knowledge and would also need to be repeated for any new dataset, or if risk factors change over time.

Previous research using automated methods for HIE model development with EHRs collected before delivery obtained AUCs of 0.87 (95% CI 0.86, 0.88) with logistic Elastic net^29^, AUC of 0.78 (95% CI 0.77, 0.79) using deep learning^30^ and 0.72 AUC (95% CI 0.71, 0.73) with logistic ridge regression^30^. While the latter results are in line with findings of this study, data were collected over different time periods and cannot be compared easily.

Our strategy for model development shared similarities with a recent published protocol for estimating risk of acute kidney injury using EHRs^14^, including data quality control, defined participant inclusion/exclusion criteria, separate training and test data to avoid overfitting, feature engineering such as creating dummy variables from unordered categorical variables, use of model performance metrics that balance sensitivity with specificity and comparison of multiple model architectures. Assessment of calibration was not suitable for this data given its age. The CPP data lacked sufficient granularity for effective application of recurrent neural network designs which make use of temporal relationships between features^14^ and are ideal for real-time updated risk stratification using data collected throughout the patient pathway. Instead, our models were trained using data equivalent to measures obtained during clinic assessment but do demonstrate feasibility with this different type of data.

Although our results do not support a significant improvement in HIE discrimination following the addition of intrapartum or birthweight variables, our findings illustrate the possibility of updating risk estimation using data collected at different time points throughout patient care as has been done in real-time for acute kidney injury demonstrating the potential for this approach^14^. A key concern with automated model development is the potential for classifiers to learn from physician behaviour and beliefs rather than provide new diagnostic insight of disease process^31^. Predictions based on physicians’ behaviour are unlikely to provide improvements in individual-level care beyond the average action taken for an average patient^31^. These challenges also apply in perinatal prediction, for example women with risk factors for stillbirth are delivered earlier and therefore prediction of stillbirth after 37 weeks will be ineffective^32^. To ensure models are unaffected by these biases, extensive external validation in different clinical environments where policies and procedures differ is needed.

While automated and manual feature selection processes gave equivalent classification performance, the automated approaches required up to three-times more features (n=60 vs n=20-35) suggesting some redundancy that clinical insight can circumvent. Simpler models are more desirable because fewer variables are required to obtain a risk estimate and have wider utility in the presence of missing data. Overfitting is also a concern when the number of features is large compared with the sample size which can produce inflated coefficients and imprecise standard errors resulting in poor generalisability and model instability^33^. Provided appropriate external validation and quality control procedures are in place these limitations should not be a barrier to clinical utility.

Using top Elastic net feature rankings, we explored a range of classifiers capable of modelling non-linear relationships and interactions but found no strong improvement in classification performance compared with logistic regression. These results were anticipated as the data were mostly categorical and linearity assumptions were justified. Conditioning on linearity and residual homogeneity through feature selection with Elastic net could also explain a lack of improvement. These findings are consistent with other studies^34,35^ and highlight that logistic regression is well suited for automated risk estimation using this type of data. Regression models are also desirable for providing well-calibrated risk estimates that are easy to interpret and already familiar with clinicians in contrast with non-parametric methods.

We used a dataset collected in the 1950s and our specific findings on HIE prediction may not translate into current clinical practise. Studies of real-time updated risk stratification applied to pregnancy outcomes in contemporary data are needed. Although the dataset was large, the number of HIE cases was small creating class imbalance issues that are known to adversely affect model performance^36^. We explored subsampling of the training dataset to address this issue but found no improvement in discrimination (not shown).

This study explored the concept of real-time updated risk stratification applied to disease prediction with exemplar application to HIE using data collected through pregnancy. While we find no substantial improvement in prediction using intrapartum or birthweight in combination with antenatal factors, this study does demonstrate the feasibility of such approaches. Future studies are needed to evaluate real-time updated risk stratification of other outcomes.

## Data Availability

Stata, R and Python3 code to replicate these analyses are available from https://github.com/MRCIEU/hie-ml.
The CPP data files and documentation are available for download from the National Archives Catalog (https://www.archives.gov/research/electronic-records/nih.html)

## Data and open-source code availability

Stata, R and Python3 code to replicate these analyses are available from https://github.com/MRCIEU/hie-ml.

The CPP data files and documentation are available for download from the National Archives Catalog (https://www.archives.gov/research/electronic-records/nih.html)

---

**Supplementary Table 1.** Established risk factors for HIE.

| Antenatal Factors (n=20) | Intrapartum Factors (n=14) | Infant birthweight (n=1) |
| --- | --- | --- |
| - Maternal age (<20, 20-24, 25-29, 30-34, >35)<br>- Parity 0, 1,>1<br>- Maternal Employment<br>- Health Insurance<br>- Maternal race<br>- FHx of seizures (recurrent non-febrile seizures)<br>- FHx of neurological disorder (excludes seizures)<br>- Infertility Treatment<br>- Maternal Hypertension<br>- Maternal height (<160, 160-164, >164)<br>- Maternal Thyroid Disease<br>- Pre-eclampsia<br>- Antenatal bleeding (mod or severe)<br>- Viral Illness<br>- Alcohol (some, none, unknown)<br>- Birthweight centile (>90<sup>th</sup>, 10-90<sup>th</sup>, 3<sup>rd</sup>-9<sup>th</sup>, <3<sup>rd</sup>)<br>- Sex<br>- Abnormal placenta<br>- Late or no antenatal care<br>- Multiple births | - Gestation (37-42)<br>- OP presentation<br>- Maternal Pyrexia<br>- Maternal Intrapartum Event (Haemorrhage, convulsions, uterine rupture, snapped cord, out of hospital birth)<br>- Membrane rupture >12 hours<br>- Blood Pressure abnormalities – Captured above<br>- Nuchal cord<br>- Cord prolapse<br>- Onset of labour (spontaneous, induced, none)<br>- Mode of delivery (Spontaneous, induced vaginal, elective CS, emergency CS, breech manoeuvre)<br>- Shoulder dystocia<br>- Epidural Anaesthetic<br>- Breech Presentation<br>- ROM>12 hours | - Birthweight centile (>90<sup>th</sup>, 10-90<sup>th</sup>, 3<sup>rd</sup>-9<sup>th</sup>, <3<sup>rd</sup>) |

Previously reported predictors of hypoxic-ischaemic encephalopathy obtained from Badawi
*et al*<sup>19,20</sup>. FHx, family history. OP, occiput posterior fetal position. CS, caesarean section.
ROM, rupture of membranes.

**Supplementary Table 2.** Feature selection methods.

| Method | Theory of operation |
| --- | --- |
| LASSO regression | Applies an L1 penalty term that shrinks weak predictors to zero such that they are removed from the regression model<sup>10</sup>. Feature importance is estimated using the absolute coefficient. |
| Elastic net regression | Applies L1 and L2 penalty terms which both downweights and removes weak predictors from the regression model<sup>11</sup>. Feature importance is estimated using the absolute coefficient. |
| Random forest | An ensemble of decision trees. Feature importance is estimated using Gini impurity metric which measures the probability of an incorrect classification for each feature<sup>37</sup>. |
| Linear support vector classifier | Applies an L1 penalty term that shrinks weak predictors such that they are removed from the support vector classifier (SVC)<sup>10</sup>. The SVC learns a decision boundary around features for each class. |
| Recursive feature elimination | Backward stepwise logistic regression that compares model performance using the area under the ROC curve<sup>13</sup>. |

LASSO, least absolute shrinkage and selection operator. L1 penalty, shrinks weak coefficients to zero. L2 penalty, down weights weak coefficients in the model.

**Supplementary Figure 1.**
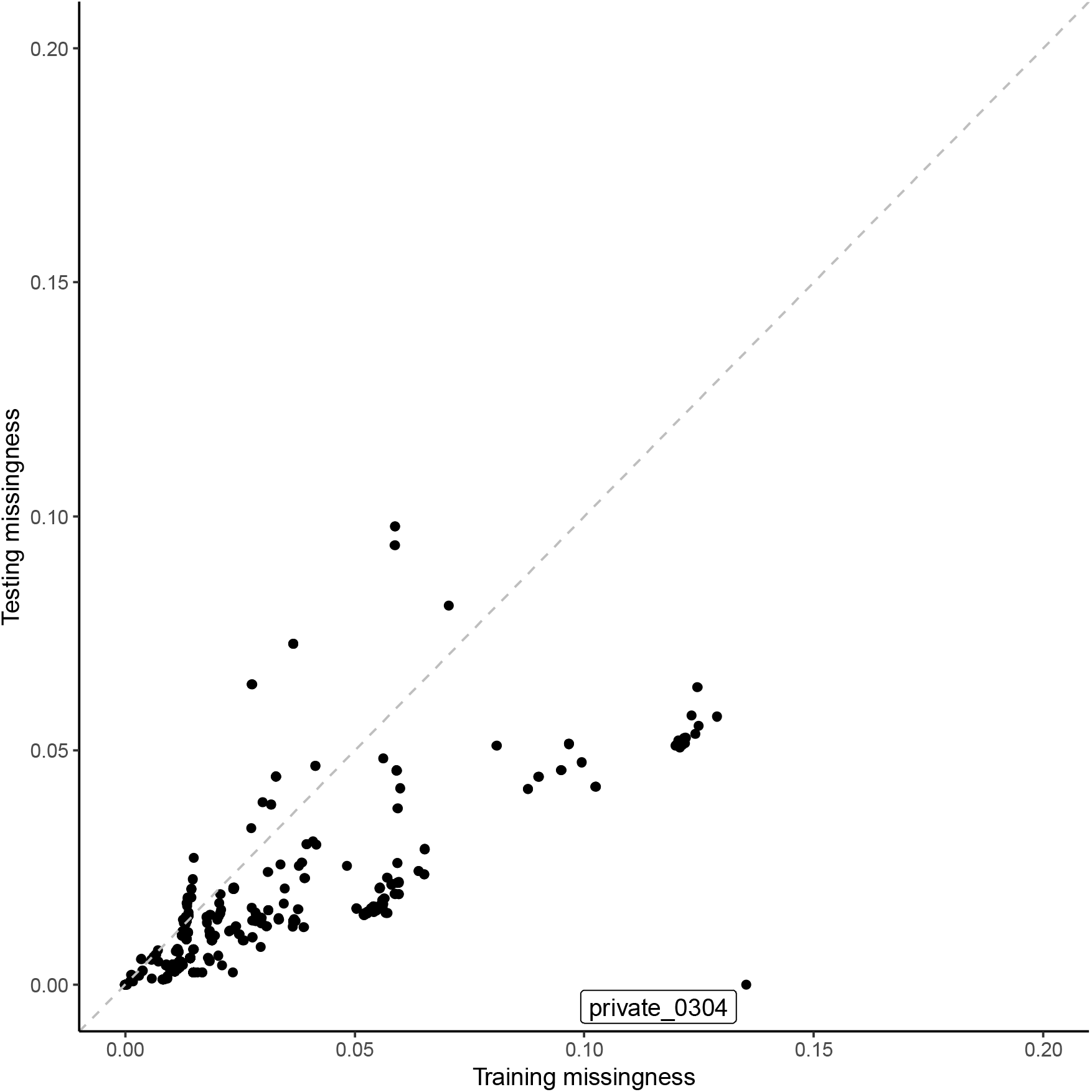
Variable missingness in testing and training datasets. Proportion of variable missingness in testing and training datasets. Variables with an absolute difference in missingness greater than 10% were labelled. Private_0304, private health insurance.

**Supplementary Figure 2.**
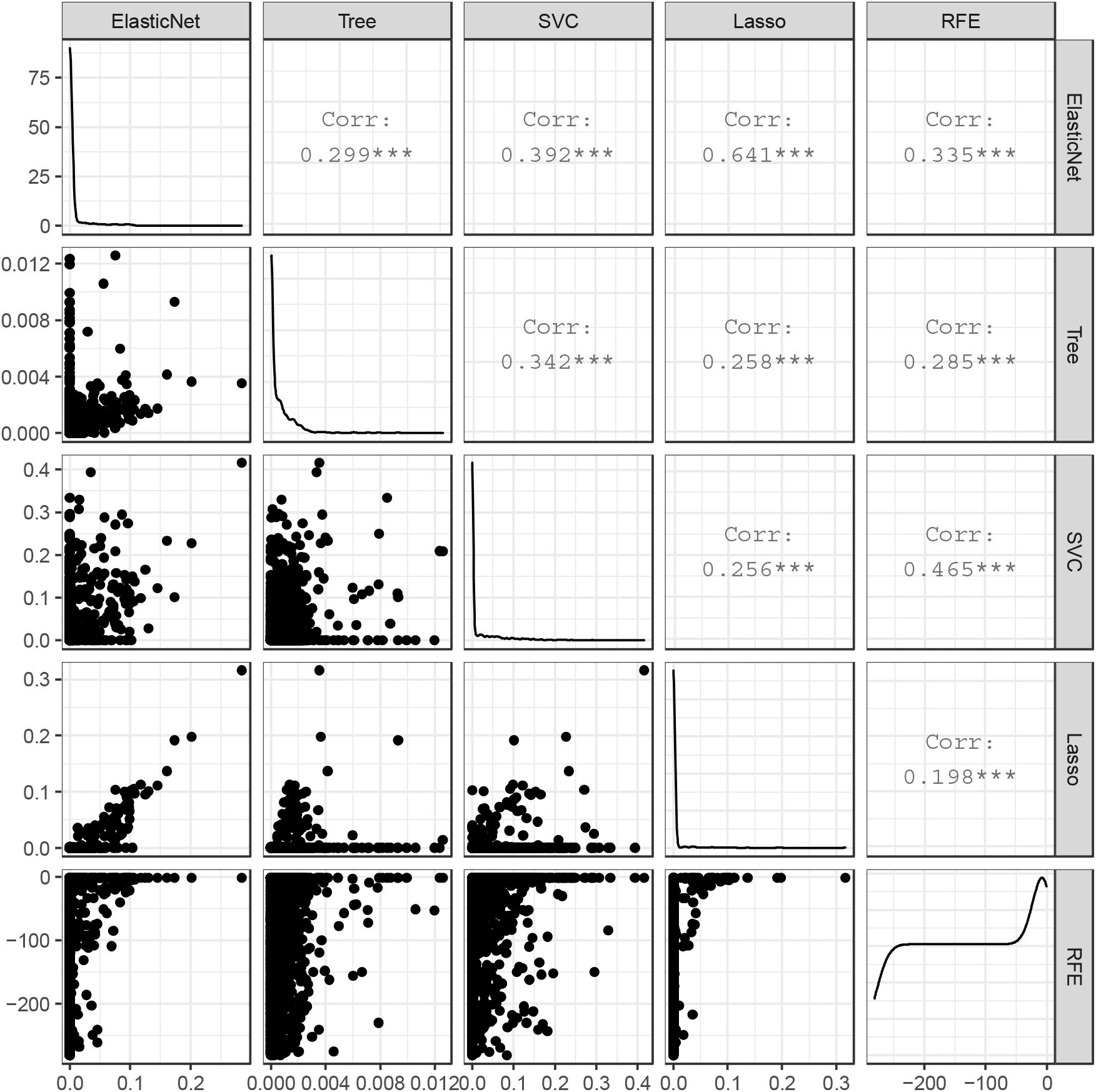
Distribution and correlation of automated feature selection scores using antenatal features. L1 penalised regression (Elastic net & LASSO), absolute regression coefficient. Random forest classifier (Tree), feature importance. L1 penalised linear support vector classifier (SVC), absolute coefficient. Inverse reverse feature elimination (RFE) ranking using logistic regression. Corr, Spearman’s rho rank correlation coefficient.

**Supplementary Figure 3.**
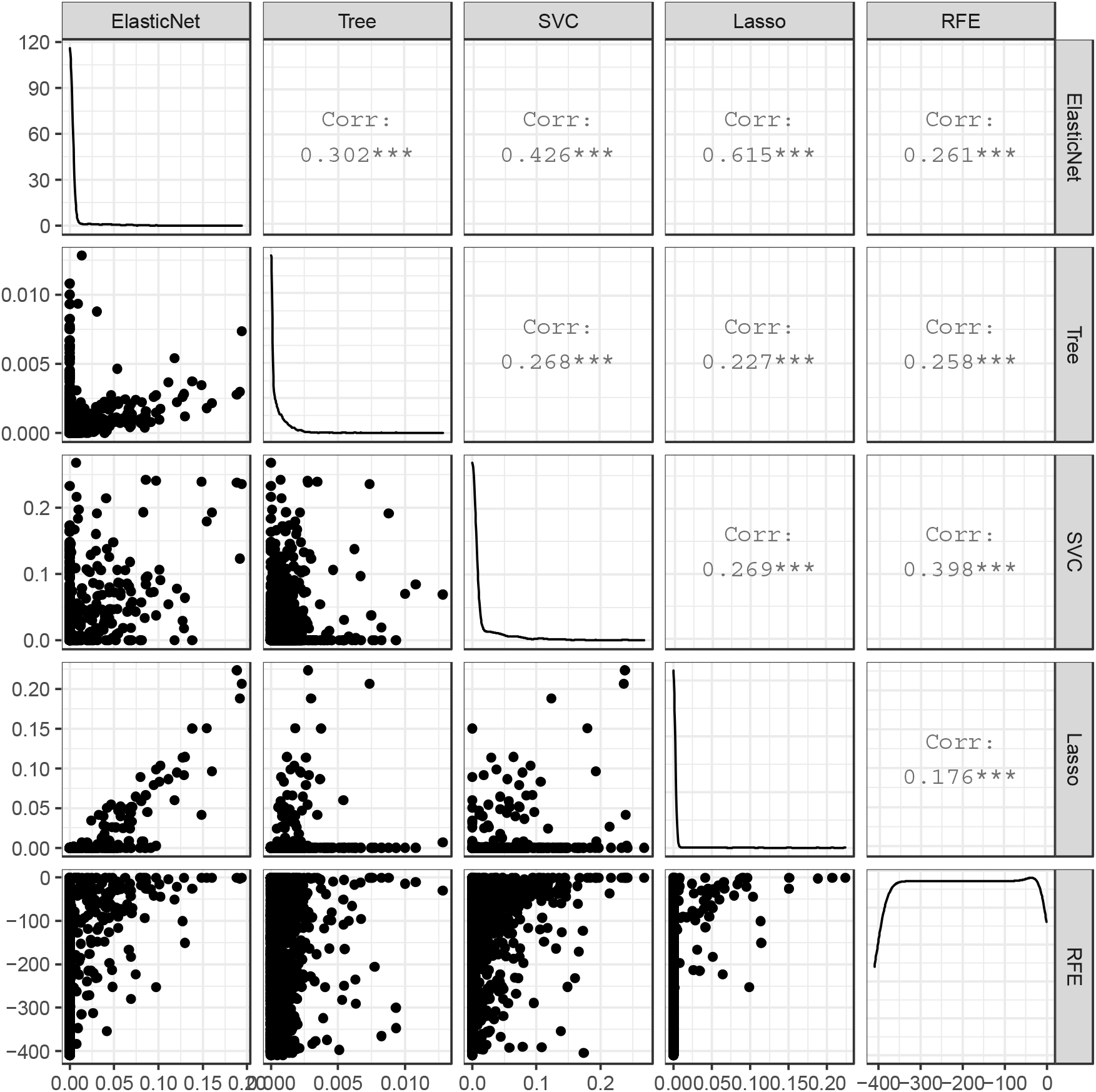
Distribution and correlation of automated feature selection scores using antenatal and intrapartum feature set. L1 penalised regression (Elastic net & LASSO), absolute regression coefficient. Random forest classifier (Tree), feature importance. L1 penalised linear support vector classifier (SVC), absolute coefficient. Inverse reverse feature elimination (RFE) ranking using logistic regression. Corr, Spearman’s rho rank correlation coefficient.

**Supplementary Figure 4.**
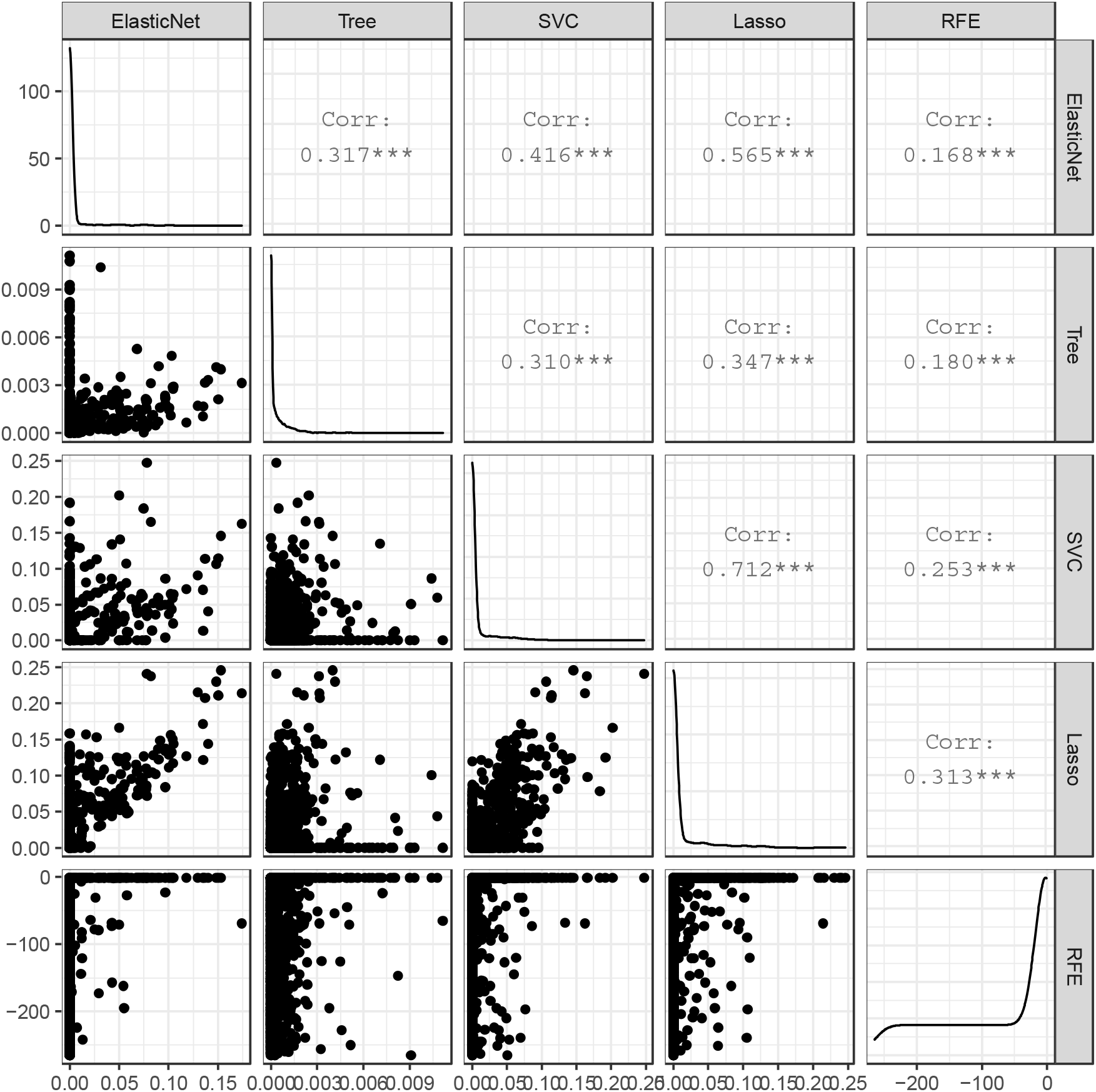
Distribution and correlation of automated feature selection scores using antenatal, intrapartum and birthweight feature set. L1 penalised regression (Elastic net & LASSO), absolute regression coefficient. Random forest classifier (Tree), feature importance. L1 penalised linear support vector classifier (SVC), absolute coefficient. Inverse reverse feature elimination (RFE) ranking using logistic regression. Corr, Spearman’s rho rank correlation coefficient.

**Supplementary Figure 5.**
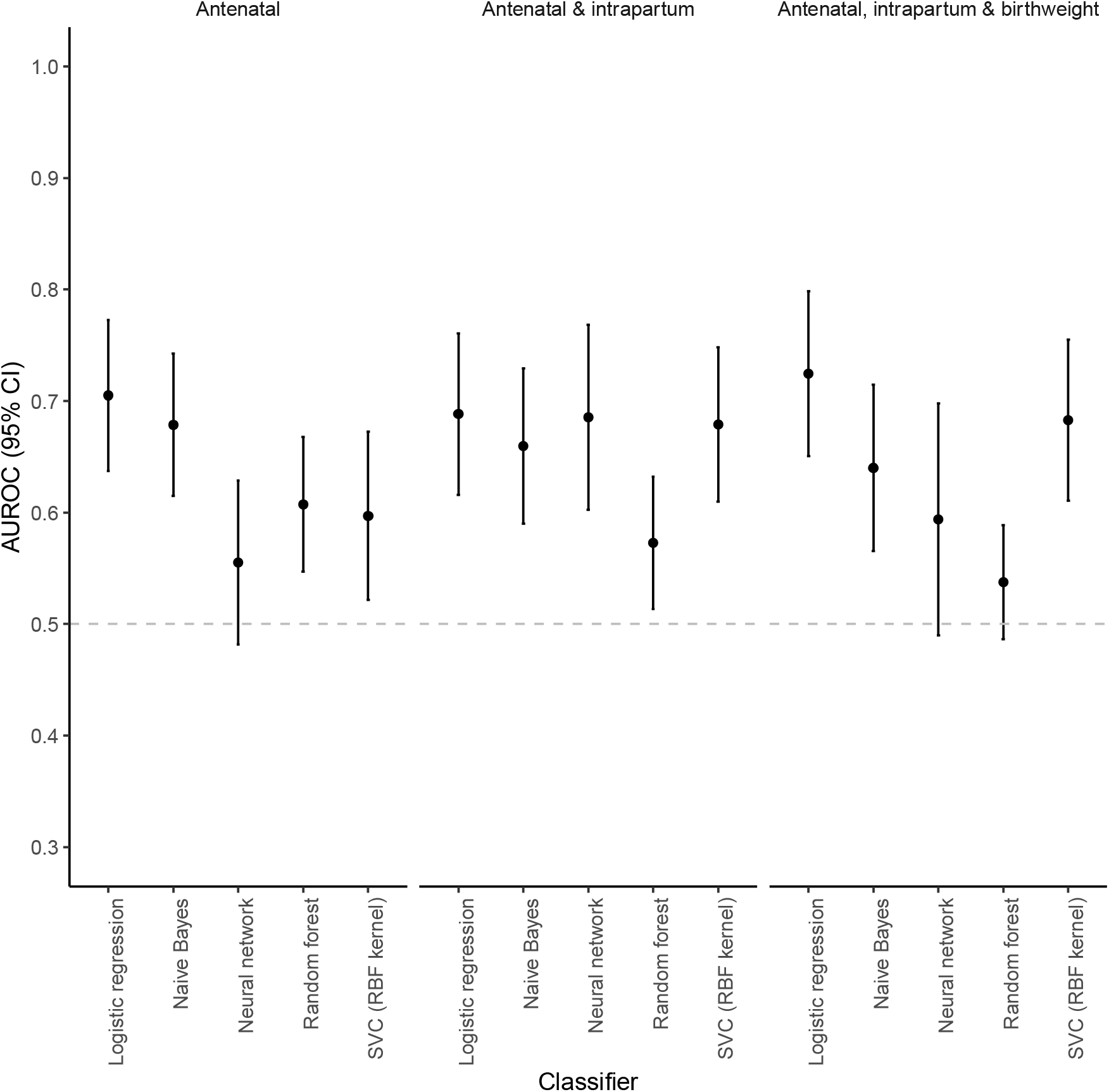
Discrimination of HIE with a range of classifiers. Sixty features selected using logistic Elastic net from the antenatal, intrapartum and birthweight dataset. AUROC, area under the receiver-operator curve. CI, confidence interval. SVC, support vector machine. RBF, radial basis function.

